# Process evaluation protocol for the SLEEP study: a hybrid digital CBT-I intervention for working people with insomnia

**DOI:** 10.1101/2022.03.03.22271613

**Authors:** A. Hurley-Wallace, T. Moukhtarian, K. Patel, C. Toro, S. Russel, G. Daley, L. Walasek, N. K. Y. Tang, C. Meyer

## Abstract

**Introduction:** The Supporting Employees with Insomnia and Emotional Regulation Problems (SLEEP) pilot uses hybrid digital Cognitive Behavioural Therapy for Insomnia (CBT-I) to help working people address their sleep and emotion regulation problems, using cognitive, behavioural, and psychoeducation techniques. A process evaluation within this trial will improve our understanding of how the intervention brought about change, including identifying barriers and facilitators to engagement and subsequent change, and the extent to which contextual factors played a role.

**Methods and analysis:** This qualitative process evaluation will use semi-structured interviews, conducted via online videoconferencing, to explore participant experiences of the SLEEP intervention. Twenty-five participants who completed the SLEEP intervention (16 %) will be randomly sampled. Interviews will be analysed using a thematic and framework analytic approach. A codebook style thematic analysis will be used, and a framework focusing on the research questions will be applied to the codebook. The final report will present themes generated, alongside the finalised codebook. The report resulting from this research will adhere to CORE-Q quality guidelines.

**Ethics and dissemination:** Full approval for the SLEEP study was given by the University of Warwick Biomedical and Research Ethics Committee (BSREC 45/20-21). All data collection adheres to data collection guidelines. Participants provided written informed consent for the main trial, and all interviewees provided additional written and verbal (audio-recorded) consent. The results of the process evaluation will be published in a peer-reviewed journal and presented at conferences. A lay report will be provided to all participants.

**Trial registration:** ISRCTN13596153; Pre-results.

---

The Supporting Employees with Insomnia and Emotional Regulation Problems (SLEEP) study is one of six studies^1^ being conducted as part of the Midlands Engine Mental Health and Productivity Pilot (<u>mhpp.me</u>). This group of studies targets mental health and productivity in the working population, by engaging with employer stakeholders throughout the Midlands, UK. SLEEP is a hybrid, digital Cognitive Behaviour Therapy for Insomnia (CBT-I) and emotion regulation (ER) intervention. The protocol for the SLEEP trial (a randomised waitlist-control trial) is reported elsewhere.[1] This protocol describes the process evaluation of the hybrid intervention only.

Established diagnostic systems define insomnia as dissatisfaction with sleep quantity or quality, manifested as difficulty initiating sleep or maintaining being asleep, present for at least three nights per week over three months.[2, 3] Chronic insomnia symptoms are reported by 30-48% of adults in the general population.[4, 5] Insomnia can impact functioning in social, psychological, educational, and occupational domains, resulting in significant economic burden to healthcare systems.[6, 7] In the occupational domain, poor sleep has been related to increased absenteeism, reduced productivity, and poorer work satisfaction.[6, 8] Thus, insomnia represents a major public health concern with substantial occupational health risks to both shift and non-shift workers.[9, 10] RAND Europe reports that one in three workers in the UK are affected by sleep problems, costing the UK economy approximately £36 billion every year, due to productivity loss.[11]

Resources for employees to improve sleep are limited in workplaces, despite evidence that they have the potential to improve productivity and reduce absenteeism.[12] CBT-I is the recommended first-line treatment for insomnia.[13, 14] Meta-analyses have estimated a moderate to large positive effect of CBT-I on sleep efficiency and quality in individuals with insomnia.[15] A 2020 systematic review also identified two studies that have evaluated the effectiveness of CBT-I for employees,[16-18] where one study found improvements in work productivity.[18] Given the number of working people with sleep problems, delivering digital CBT-I through workplace settings has the potential to increase accessibility, whilst also improving the cost-effectiveness of treatment delivery.[19] Recent cost-benefit analyses of digitised CBT found cost savings of between £116-£136 million per annum in England compared to face-to-face therapy.[20]

Sleep problems are also closely related to difficulties people may experience in regulating their emotions,[21] where research indicates that people with insomnia are five-times more likely to report anxiety and depression symptoms.[22] Hence, there is a need for hybrid interventions addressing insomnia and ER issues. To our knowledge, no such interventions have been trialled with employees in workplace settings. Hybrid treatment approaches have been piloted in the management of health conditions such as chronic pain, with promising improvements in sleep outcomes, mood, and fatigue.[23]

The SLEEP study adopts a hybrid, digital CBT-I+ER approach. The protocol of the SLEEP trial is freely available [1]. Briefly, the study aims to evaluate the effectiveness of the SLEEP intervention for employees and self-employed working people in the Midlands region. The

Midlands region of the UK has a population of approximately 11 million, with 4.5 million jobs.[24] Participants are asked to fill out a sleep diary over 6-weeks, and are suggested to spend around 1-hour each week working through content on the digital platform (Table 1). In addition, they are offered four 1-hour videoconferencing sessions with therapists trained specifically for the SLEEP trial. Primary outcomes include insomnia severity, and depression and anxiety symptoms. Secondary outcomes include sleep pattern, mental wellbeing, quality of life, job satisfaction and work productivity.

**Table 1.** Summary of SLEEP intervention content based on CBT-I [25, 26] and ER manual [27]components.

| Week number | Intervention components |
| --- | --- |
| 0 | • Introduction of the SLEEP diary |
| 1 | • Sleep and mental health<br>• CBT formulation, including vicious cycles of insomnia<br>• Sleep science and psychoeducation<br>• Sleep hygiene psychoeducation |
| 2 | • **Online therapist contact: session 1**<br>• Review of sleep diary<br>• Set up new sleep plan<br>• Importance of sticking to the programme<br>• Sleep psychoeducation: Predisposing, precipitating and perpetuating factors of insomnia<br>• Sleep restriction therapy (SRT)<br>• Stimulus control therapy (SCT) |
| 3 | • Sleep psychoeducation: sleep needs at different stages of life, chronotypes<br>• Emotion regulation (ER): Psychoeducation in physiology of stress and response to stress<br>• Emotion regulation (ER): Skills training in relaxation techniques (progressive muscle relaxation, tactical breathing, guided imagery, grounding)<br>• Sleep restriction therapy (SRT)<br>• Stimulus control therapy (SCT) |
| 4 | • **Online therapist contact: session 2**<br>• Review of sleep diary<br>• Troubleshoot sleep plan<br>• Sleep restriction therapy (SRT)<br>• Stimulus control therapy (SCT)<br>• Cognitions/ thoughts and how we understand them within the CBT model<br>• Unhelpful thinking styles and cognitive distortions<br>• Cognitive restructuring/reframing<br>• Emotion regulation (ER): Modifying emotions by managing stress/ worrying with problem solving techniques |
| 5 | • **Online therapist contact: session 3**<br>• Review of sleep diary<br>• Troubleshoot sleep plan<br>• Sleep restriction therapy (SRT)<br>• Stimulus control therapy (SCT)<br>• Healthy lifestyle choices (eating, physical activity)<br>• Work-life balance<br>• Efficient time management skills (e.g., SMART goal setting) |
| 6 | • **Online therapist contact: session 4**<br>• Review of sleep diary<br>• Troubleshoot sleep plan<br>• Demonstrating progress with visual graphs<br>• Emotion regulation (ER): Self-compassion and resilience<br>• Relapse management |

## Process evaluation

This protocol outlines a qualitative process evaluation of the SLEEP intervention trial, described above. Interviews will be conducted with a random selection of individuals who took part in the trial. Interview data will be analysed using codebook style thematic and framework analysis.[28-30]

This qualitative-focused enquiry is appropriate at this stage of intervention development, where the results of the SLEEP trial will be utilised to inform future scaling of the intervention.[31] Thematic analysis will be used to explore participant experiences, including barriers, facilitators, and contextual factors impacting engagement. At this stage, as well as exploring participant experiences, it is important to consider the underlying behavioural mechanisms that may underpin how the intervention itself impacts outcomes. To do this, participant feedback will be mapped to the Behaviour Change Techniques (BCTs) Taxonomy.[32] Identified BCTs will be presented separately to the thematic analysis in the process evaluation report. In summary, the process evaluation aims to:

1. Explore participants’ experiences of the intervention, including facilitators and barriers to engagement with the intervention, and any unexpected benefits and/ or consequences of the intervention.
2. Identify to what extent contextual factors (e.g., varied working and living circumstances, changing context of the COVID-19 pandemic) may have impacted outcomes of the intervention, focusing on outcomes relating to sleep and emotional regulation.
3. Identify potential behaviour change mechanisms that may contribute to intervention outcomes, by drawing from participant feedback from interviews.

## Methods

The process evaluation study report will adhere to the CORE-Q reporting guidelines for qualitative research.[33]

### Interviews

This study will use semi-structured, individual (1:1) interviews conducted via videoconference (Microsoft Teams) to collect qualitative data. The interview schedule, consisting of 8 questions, and several prompts, is provided in Table 2. Data analysis will use codebook style thematic and framework analysis.[28-30] This approach is selected for the process evaluation for two key reasons: (i) this set of interviews will be conducted by six members of research team, and analyses will seek to incorporate the interpretations of all interviewers; one primary researcher will, however, be responsible for the final interpretative decisions pertaining to the codebook and themes, and (ii) the process evaluation seeks to explore specific topics, including identifying barriers and facilitators to change, hence the study has predetermined informational needs.[34]

**Table 2.** Semi-structured interview schedule.

| Question | Prompts |
| --- | --- |
| 1. What was your overall experience of being in the programme? | What aspect of the programme did you like the most? (e.g. educational components, practical solutions/tools, therapist contact, online/digitalisation, flexibility around commitments)<br>Why did you like *[repeat back answer]*?<br>What aspect of the programme did you like the least? (same prompts as above)<br>Why didn't you like *[repeat back answer]*? |
| 2. What did you think about the format in which the programme was delivered? (e.g. length, resources, content) | How was your experience with the digital platform/online content? (e.g. usability/ user-friendly/ easy to use?)<br>How was your experience with your therapist?<br>What did you think about the topics covered during the online therapy sessions?<br>Did you feel there were a suitable number of therapy sessions?<br>What do you think about how long the programme lasted? (too long/ too short/ the right length)<br>What do you think about the online/digital nature of |
|  | the programme? |
| 3. How did you feel that the programme was tailored to your needs? | General prompts – e.g. can you tell me more about that?/<br>Can you expand on that? |
| 4. What were your initial expectations of the programme, if any? | Did the programme meet your expectations?<br>What impact did you anticipate the programme would have on your <u>sleep</u>, and did it meet those expectations?<br>Can you tell me about any changes to your <u>sleep</u> that you have experienced? |
| 5. What impact did you anticipate the programme would have on your <u>physical or mental wellbeing</u>, and did it meet those expectations? | Can you tell me about any changes to your <u>physical or mental wellbeing</u> that you have experienced?<br>As a result of the programme, did you experience any changes in other areas of your life? (e.g. social, work-life balance) |
| 6. Were there any barriers that you experienced during your participation in the programme? | Were you able to complete the required activities around your existing commitments? (e.g. attend appointments, complete online content)<br>Did you have access to a private space and/or necessary IT equipment? |
| 7. Did you have any concerns about stigma during your participation in this study? (e.g. from employer, colleagues, family) | If so, can you tell me more about this? |
| 8. If you had the opportunity, what would you most like to change about the programme? | Is there anything that you wish had been in the programme, but was not there? |

### Research team description

The team of researchers conducting interviews and involved in subsequent qualitative analyses consists of six researchers with academic and/or clinical backgrounds in psychology. Three members of the team hold PhDs and three members of the team hold masters-level qualifications in psychology. The primary researcher (AHW) holds a PhD with specialisation in health psychology. AHW’s epistemological stance is critical realist. The extent to which individual team members’ epistemological and ontological stances influenced qualitative findings will be discussed in the study report. Two members of the research team had no prior relation to the study participants. Four members of the research team (including AHW) worked as therapists within the SLEEP intervention; therapists did not interview participants who had been their therapy clients during the programme.

### Sample size

The target sample size for this process evaluation will be 25 interviews (16% of the sample). Notably, research on saturation serves as a guide for qualitative research protocols, however, within thematic analysis new meanings are always theoretically possible.[35, 36] Research on saturation in thematic analysis indicates that saturation of meaning approximates 16 to 24 interviews and represents an in-depth understanding of identified issues.[37] Therefore, interview data will be added to analyses in batches, either until the data is saturated across cohorts (five cohorts), or the target of 25 interviews is fulfilled.

### Recruitment

Twenty-five participants will be randomly sampled across cohorts (five cohorts). The sample will be stratified, aiming for an equal weighting of participants between the main and wait-list control arms of the trial, and from within each cohort. Participants will be invited, by email, to take part in an interview. If they do not respond, they will be followed up by email after 1-week. If there is no response after this, no further contact will be made. Should a participant be approached and decline/ not respond to the interview invite, another participant will be randomly selected from those remaining within that same cohort.

### Consent

We will only recruit from participants who consented to take part in follow-up interviews when consenting to the SLEEP trial.[1] Interviews will require a written and verbal consent form. The consent form will be signed electronically by the interviewee and returned by email prior to attending the interview. Verbal consent will additionally be recorded, at the beginning of the interview, and signed-off individually for each participant, by the interviewer.

Audio-only recording will be used for both verbal consent and interviews. Audio will be recorded using OBS studio (obsproject.com). Microsoft Teams was not used to record consent or interviews, as video recording was not necessary, and there is no option to record audio-only on the Microsoft Teams platform.

### Procedure

Each participant will be allocated an interviewer from the research team, who will be independent from the treatment delivery team of the respective interviewee. This is to ensure confidentiality, as well as to ensure the interview setting allows participants to express their honest opinions. Participants will be invited to attend an online videoconference meeting via Microsoft Teams at a mutually agreed time (agreed by email with the research team).

Interviewees will be greeted by the interviewer informally in the first instance and the interviewer will read a standardised introduction. The interviewer will start the audio-recording, using OBS studio, and will complete the verbal consent form prior to starting the interview. The interviewer will then commence the interview, following a semi-structured, open-ended interview schedule (Table 2). Interviews are expected to last up to approximately 45-minutes. At the end of the interview, the recording will be stopped, and participants will be given an opportunity to ask any questions they may have.

Audio recordings will be saved in a secure, limited access, digital folder within the University of Warwick servers. Recordings will then be sent to a third-party University of Warwick approved vendor to be transcribed verbatim. Transcriptions will be anonymised by the external provider. Each interviewer will then cross-check their transcript(s) against the original audio-recording of their interviews(s) for accuracy.

### Qualitative analysis

A flowchart of the data analysis approach is provided in Figure 1. Data analysis will combine codebook style thematic and framework analysis.[28-30]. The underpinning foundation of this approach is interpretative.[28, 34] There are several benefits to using a codebook style approach for thematic analysis, including:

**Figure 1.**
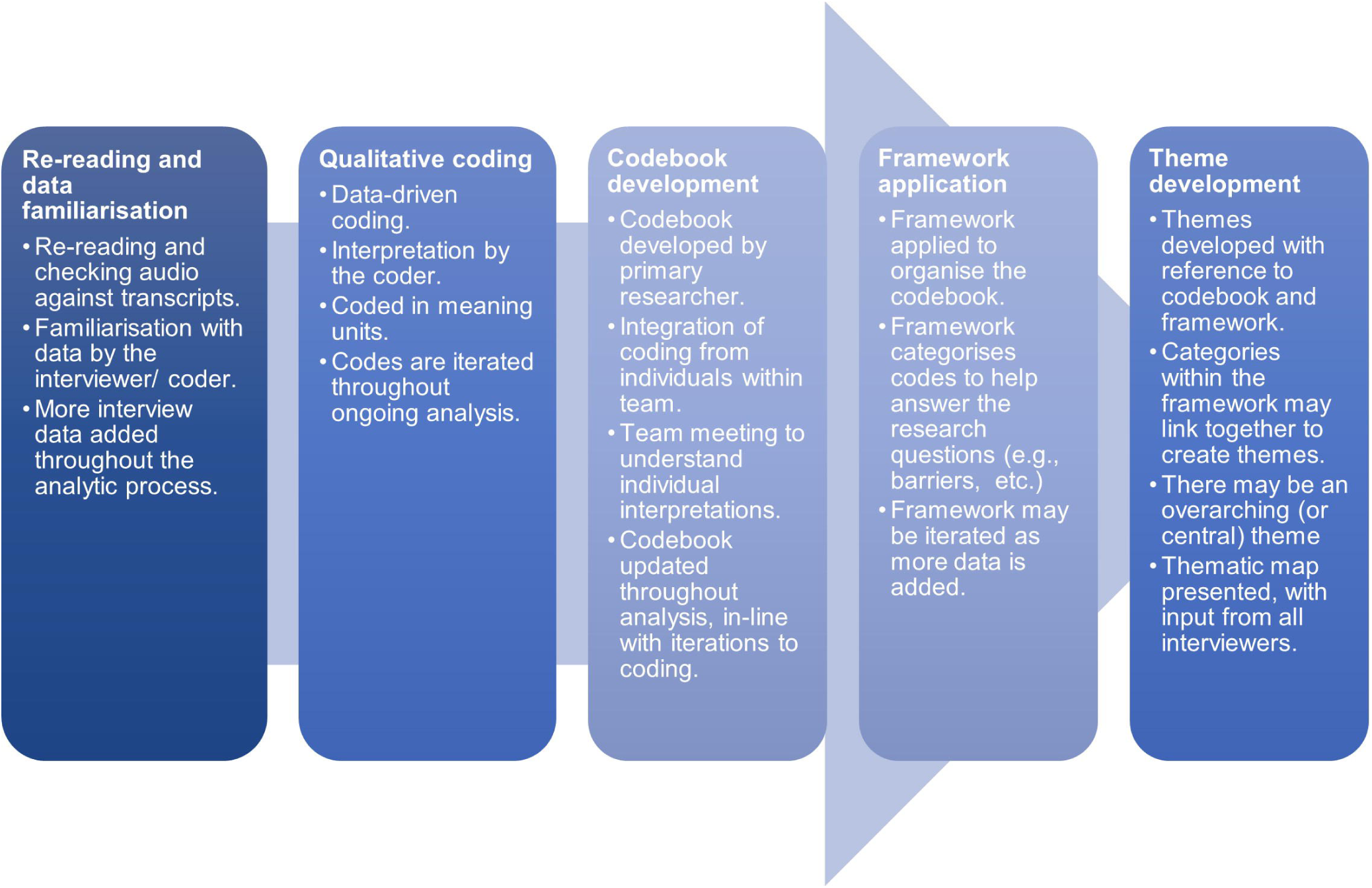
Flowchart of the analytic approach; incorporating codebook style thematic, and framework analysis.

- Facilitating the exploration of specific topics, in cases where qualitative research has predetermined informational needs.[34]
- Utilising several team members to increase the efficiency of the analysis, without sacrificing the interpretative underpinnings of thematic analysis (as can happen in ‘coding reliability’ approaches).[28]
- Enabling team members who are not experienced with qualitative analyses to gain experience, with guidance from more experienced team members (at least one experienced qualitative lead needed).
- Encouraging triangulation of ideas throughout analyses, where the process naturally incorporates different individual’s perspectives.

Following the six phases of thematic analysis,[38] qualitative data collected in this study will be initially reviewed by each interviewer, using original audio-recordings to cross-check transcriptions. Once transcriptions have been checked and edited as needed, they will be imported to NVivo [39] for analysis. Initial qualitative coding of the first six transcriptions will be completed by the interviewing researcher, based on their interpretation. Coding will be conducted in meaning units (anything from a phrase to a paragraph), and some coding overlap (2 to 3 codes) is allowed. Once the first six interviews have been coded, the interviewers will meet to discuss their interpretations and exchange ideas.

The primary researcher (AHW) will then collate these codes into a codebook (full set of codes with descriptions). This will involve iterations of codes, including combining similar or overlapping codes as necessary. This codebook will then be used to help chart the developing analysis.[34] A framework based on the research questions we are seeking to answer (i.e., participants experiences of the intervention; barriers and facilitators to engagement, contextual factors impacting change) will be applied to help organise the codebook.[40]

Once the codebook and framework have been initially defined, the remaining interview transcripts will be added into the analysis, using this codebook as a guide for coding new data (Figure 1). At this stage, interviews may be coded by any member of the analytic team, and as such the codebook is passed between team members. New codes can be added as needed, as qualitative coding remains a data-driven process. Both the codebook and framework may be iterated as more interview data is incorporated; final decisions on iterations to the codebook will be made by the primary researcher (AHW). Themes will be generated once all interview data has been coded, to a point of saturation across cohorts [37], and the codebook has been finalised.

The qualitative team (including all interviewers) will then meet to discuss themes and conceptualise a thematic map to fit the interview dataset. Themes will additionally be discussed and triangulated with the SLEEP study’s lead clinical psychologist (NT). Finalised themes will be presented in the process evaluation research report, with anonymised quotes to demonstrate claims. The finalised codebook will also be appended to the research report.

### Identifying potential behaviour change mechanisms

The process evaluation for the SLEEP study will also consider the underlying behavioural mechanisms that may have impacted intervention outcomes (i.e., changes to sleep and emotion regulation). Hence, the process evaluation report will, separately from the thematic analysis and thematic map produced, seek to identify potential BCTs,[32] based on participant feedback from interviews. Identified BCTs will be presented in a summary table or diagram.

### Data protection and confidentiality

All research must comply with the requirements of the General Data Protection Regulation (GDPR) and the Data Protection Act 2018. All data for this study will be electronic only on password protected drives accessible by the research team only. Audio recordings and transcriptions will be held in a digitally secure folder within the University of Warwick, with restricted access.

Audio recordings and interview consent forms will be deleted/ destroyed after analyses have been completed. Pseudonymised transcriptions will be saved in password protected word documents and will be stored on the University of Warwick shared secure servers for 10-years as per the University of Warwick regulations for data usage and storage.

### Patient and Public Involvement

This process evaluation is part of the SLEEP study which sourced patient and public involvement (PPI) input for its design. This is described in full elsewhere.[1] The process evaluation interviews did not receive input from PPI contributors.

### Trial status

Recruitment for the main SLEEP trial commenced on 18^th^ June 2021 until 31^st^ October 2021. Recruitment of participants from the SLEEP trial for process evaluation interviews is currently ongoing. Qualitative analyses are currently ongoing, where the first 10 transcripts (cohorts 1 to 4) have been coded. Interview data will be added to analyses in batches until the data across cohorts (5 cohorts) is saturated, or the target of 25 interviews is met.

### Ethics and Dissemination

The SLEEP trial and qualitative process evaluation have been granted full ethical approval by the University of Warwick Biomedical and Research Ethics Committee (BSREC 45/20-21). All data collection adheres to data collection guidelines. Participants provided written informed consent for the main trial, and all interviewees provided additional written and verbal (recorded) informed consent. The results of the process evaluation will be published in a peer-reviewed journal and presented at conferences and webinars. Lay reports will also be circulated to participating employers, their employees, and all study participants. Results will be published in an open access academic journal. Participants will not be identifiable from the study report. All quotations will be pseudonymised (names of people, organisations, and any other potentially identifying information, will be removed).

## Data Availability

All data produced in the present study are available upon reasonable request to the authors.

## Data Availability

All data produced in the present study are available upon reasonable request to the authors.

## Contributorship statement

All authors contributed to the conception, critical revision, and final approval of this manuscript for publication. AHW wrote the first draft of this manuscript, with significant guidance from TM. AHW and NKYT further developed the analytic method described, with contribution from KP regarding integration of theory. All authors are accountable for all aspects of this work.

## Footnotes

1 SLEEP (Supporting Employees with Insomnia and Emotional Regulation Problems), REST (Reducing stress in the workplace), BITE (Brief Individual Therapy for Eating disorders), PROWORK (PROmoting a sustainable and healthy return to WORK), MENTOR (Supporting employers and employees receiving treatment for Mental hEalth problems to remain eNgaged and producTive wORk) and Managing Minds.

